# Cerebrospinal fluid neurofilament light chain differentiates behavioural variant frontotemporal dementia progressors from ‘phenocopy’ non-progressors

**DOI:** 10.1101/2022.01.14.22269323

**Authors:** Dhamidhu Eratne, Michael Keem, Courtney Lewis, Matthew Kang, Mark Walterfang, Samantha Loi, Wendy Kelso, Claire Cadwallader, Samuel F Berkovic, Qiao-Xin Li, Colin L Masters, Steven Collins, Alexander Santillo, Dennis Velakoulis, The MiND Study Group

## Abstract

**Background:** Distinguishing behavioural variant frontotemporal dementia (bvFTD) from non-neurodegenerative ‘non-progressor’, ‘phenocopy’ mimics of frontal lobe dysfunction, can be one of the most challenging clinical dilemmas. A biomarker of neuronal injury, neurofilament light chain (NfL), could reduce misdiagnosis and delay.

**Methods:** Cerebrospinal fluid (CSF) NfL, amyloid beta 1-42 (AB42), total and phosphorylated tau (T-tau, P-tau) levels were examined in patients with an initial diagnosis of bvFTD. Based on follow up information, patients were categorised as Progressors. Non-Progressors were subtyped in to Phenocopy Non-Progressors (non-neurological/neurodegenerative final diagnosis), and Static Non-Progressors (static deficits, not fully explained by non-neurological/neurodegenerative causes).

**Results:** Forty-three patients were included: 20 Progressors, 23 Non-Progressors (15 Phenocopy, 8 Static), 20 controls. NfL concentrations were lower in Non-Progressors (Non-Progressors Mean, M=554pg/mL, 95%CI:[461, 675], Phenocopy Non-Progressors M=459pg/mL, 95%CI:[385, 539], Static Non-Progressors M=730pg/mL, 95%CI:[516, 940]), compared to bvFTD Progressors (M=2397pg/mL, 95%CI:[1607, 3332]). NfL distinguished Progressors from Non-Progressors with the highest accuracy (area under the curve 0.92, 90%/87% sensitivity/specificity, 86%/91% positive/negative predictive value, 88% accuracy). Static Non-Progressors tended to have higher T-tau and P-tau levels compared to Phenocopy Non-Progressors.

**Conclusion:** This study demonstrated strong diagnostic utility of CSF NfL to distinguish bvFTD from phenocopy non-progressor variants, at baseline, with high accuracy, in a real-world clinical setting. This has important clinical implications, to improve outcomes for patients and clinicians facing this challenging clinical dilemma, as well as for healthcare services, and clinical trials. Further research is required to investigate heterogeneity within the non-progressor group and potential diagnostic algorithms, and prospective studies are underway assessing plasma NfL

## Introduction

Determining the underlying cause of cognitive and behavioural change associated with functional decline is often diagnostically challenging for clinicians. Even in centres with gold-standard multidisciplinary and multimodal investigations, individuals still face several years of diagnostic uncertainty, misdiagnosis, and delay. Diagnostic delay is greatest in patients with younger onset (symptom onset before 65 years) dementia who frequently present with psychiatric or behavioural symptoms [1–4]. In particular, the behavioural, personality and executive dysfunction symptoms of behavioural variant frontotemporal dementia (bvFTD) can be difficult to distinguish from primary psychiatric symptoms and other non-neurodegenerative causes of frontal lobe dysfunction [4–12].

An increasing body of literature has identified individuals who can appear clinically to have bvFTD, meet established diagnostic criteria for bvFTD, but who do not have the expected clinical, neuropsychological and neuroimaging progression over time. Various terms and definitions have been used for group, such as “bvFTD phenocopies”, “mild behavioural impairment”, and “non-progressors”, sometimes inconsistently, and a consensus has yet to be reached [13–18]. While people with a very slowly progressive bvFTD associated with the *C9orf72* genetic mutation have been identified [19–23], it is likely that this ‘non-progressors’ group is aetiologically heterogenous, comprising people with primary psychiatric disorders or people with a mid-life stress related exacerbation of underlying personality, relationship or neurocognitive vulnerabilities. Regardless of the aetiology of phenocopy non-progressors, there is strong consensus that it is not benign, associated with high functional impact, patient distress and carer burden [13]. Differentiating between progressors and non-progressors at initial presentation, is challenging [11,15,18]. International consensus diagnostic criteria, which remain the gold standard for the diagnosis of bvFTD, involve diagnostic stratification into possible, probable or definite bvFTD [11,24]. Given that it can take years to reach a definitive diagnosis and the implications for treatment, prognosis, genetic counselling, and wider occupational and psychosocial outcomes, there is a critical need for earlier, accurate diagnostic differentiation. A biomarker that could assist in distinguishing between bvFTD progressors and non-progressors at baseline assessment could significantly improve outcomes for patients, their families, clinical trials, and healthcare systems.

Neurofilament light (NfL) is one of three subunits of a neuroaxonal cytoskeletal protein common to myelinated axons, with elevated levels in cerebrospinal fluid (CSF) and blood associated with neuronal injury and a marker of diagnosis, staging and prognosis, in a diverse range of neurological and neurodegenerative disorders [25–31]. Numerous studies have demonstrated elevated CSF and blood NfL in bvFTD compared to controls, as well as primary psychiatric disorders [29–38]. Recent expert consensus guidelines and recommendations have recommended NfL as a biomarker to assist with differentiating bvFTD from primary psychiatric disorders [11]. Our previous studies have demonstrated the ability for CSF NfL to distinguish bvFTD from primary psychiatric disorders, with high accuracy, in a real-world clinical setting, where patients who were ultimately diagnosed with a primary psychiatric illness were referred specifically for assessment of a possible neurodegenerative dementia [30] [Eratne et al., Alzheimer’s & Dementia, in press, 2021].

To our knowledge, no studies have investigated the diagnostic utility of NfL to distinguish bvFTD from non-progressor phenocopy syndromes, where there was an initial clinical suspicion for and diagnosis of bvFTD, in real-world clinical settings. The primary aim of this study was to explore the diagnostic utility of CSF NfL in differentiating progressive (neurodegenerative) bvFTD from non-progressive syndromes that presented with behavioural, psychiatric, neurological, cognitive and functional impairments, and who met diagnostic criteria for at least possible bvFTD. Secondary aims included examining differences in clinical variables between diagnostic groups, and determining the diagnostic utility of NfL and other biomarkers in differentiating between possible subtypes of non-progressors.

## Methods

This was a retrospective, longitudinal cohort study involving patients referred to Neuropsychiatry, The Royal Melbourne Hospital, a tertiary state-wide specialist service, for comprehensive multidisciplinary and multimodal diagnostic assessment of possible dementia, and ongoing management.

Inclusion criteria were: (1) A diagnosis of possible, probable or definite bvFTD [24] made at any time over the course of assessment with the service or follow-up, (2) CSF was collected as part of their diagnostic work-up between June 2009 and December 2020, and remnant CSF was available for NfL analysis, (3) available formalised follow-up information for a minimum of 12 months.

Patients were assessed in inpatient and/or outpatient settings by a multidisciplinary team including neuropsychiatrists, neurologists, neuropsychologists, nurses, occupational therapists, speech pathologists, and social workers, as well as receiving multimodal investigations including brain MRI and SPECT/FDG-PET, and CSF analysis for Alzheimer’s disease biomarkers. A consensus gold-standard diagnosis of possible, probable or definite bvFTD was made based on established diagnostic criteria [24]. With ongoing follow-up and longitudinal multidisciplinary assessments and multimodal investigations, this diagnosis could be revised as patients’ conditions changed, and as new data was made available.

Patients’ medical records were accessed, and data extracted included detailed demographic and clinical variables (Table 1), initial diagnosis, and most recent diagnosis based on serial assessments including obtaining follow up information from clinicians from other services. Structured symptom rating scales were not used consistently in clinical practice over the study period.

**Table 1.**
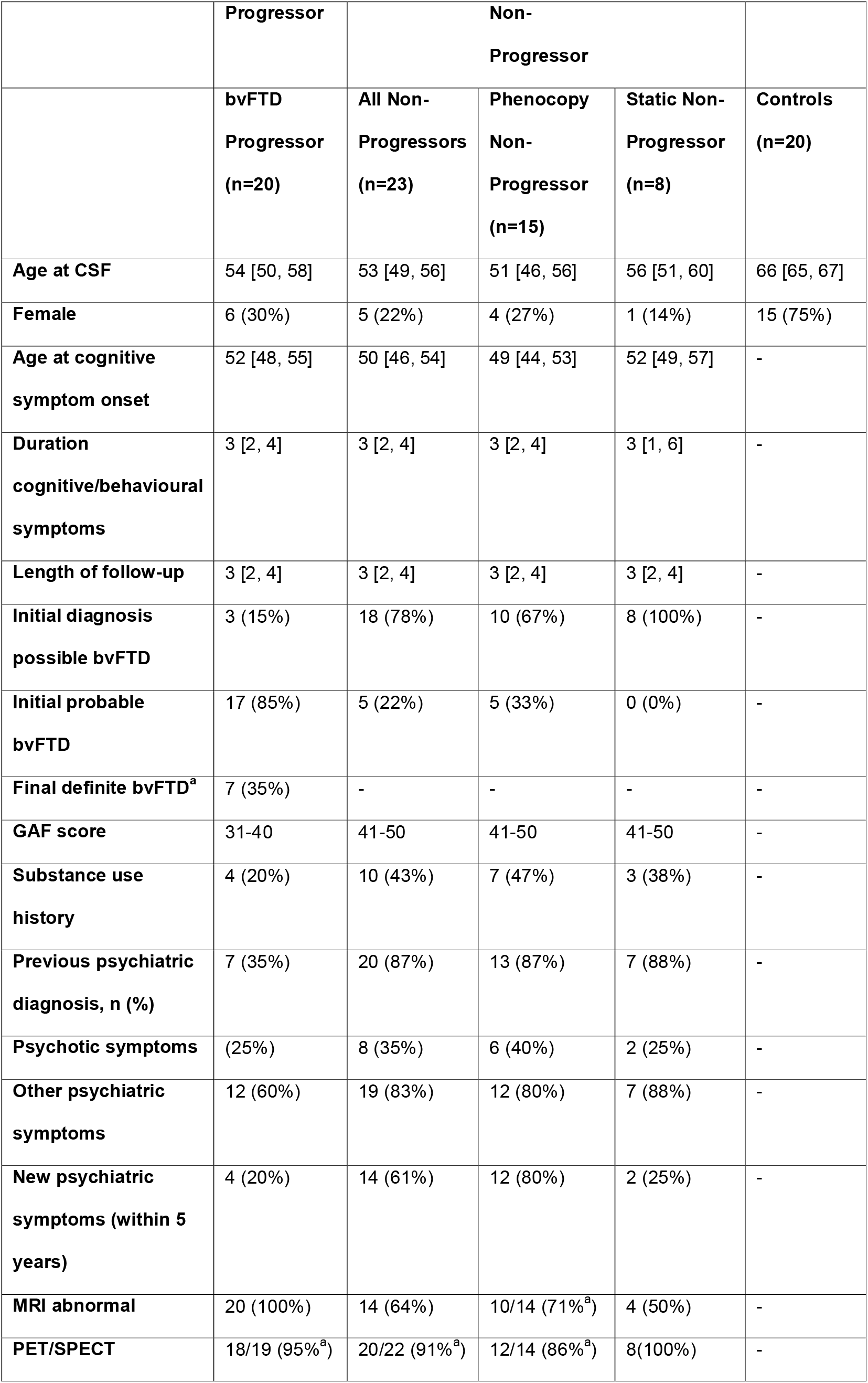
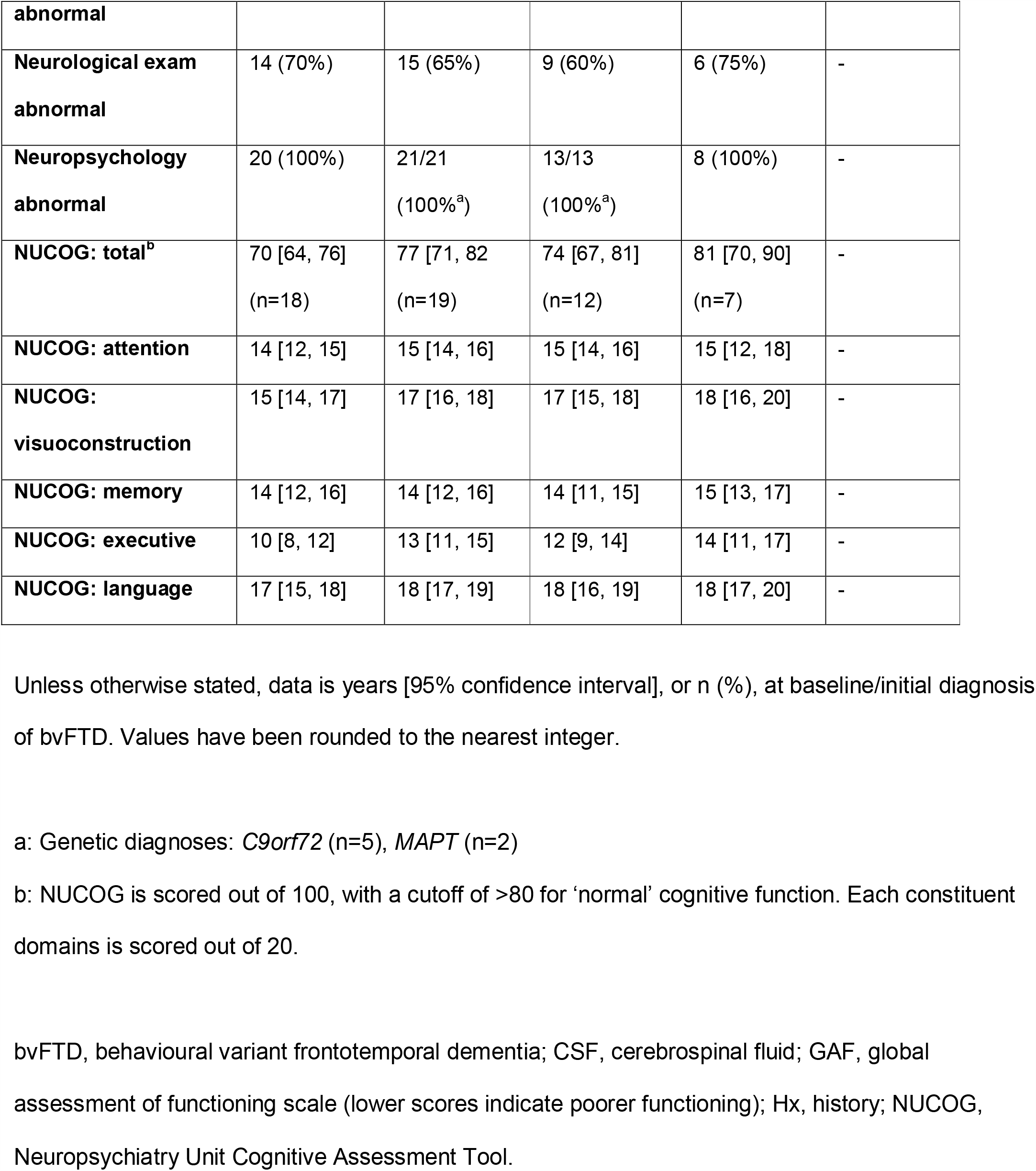
Study population demographics and clinical information.

While acknowledging some of the issues with various current terminology and definition, for the purpose of this study we elected to use terminology commonly used in the literature and in clinical practice to describe the clinical phenotypes and progression over time – Progressors and Non-Progressors (Figure 1).

**Figure 1.**
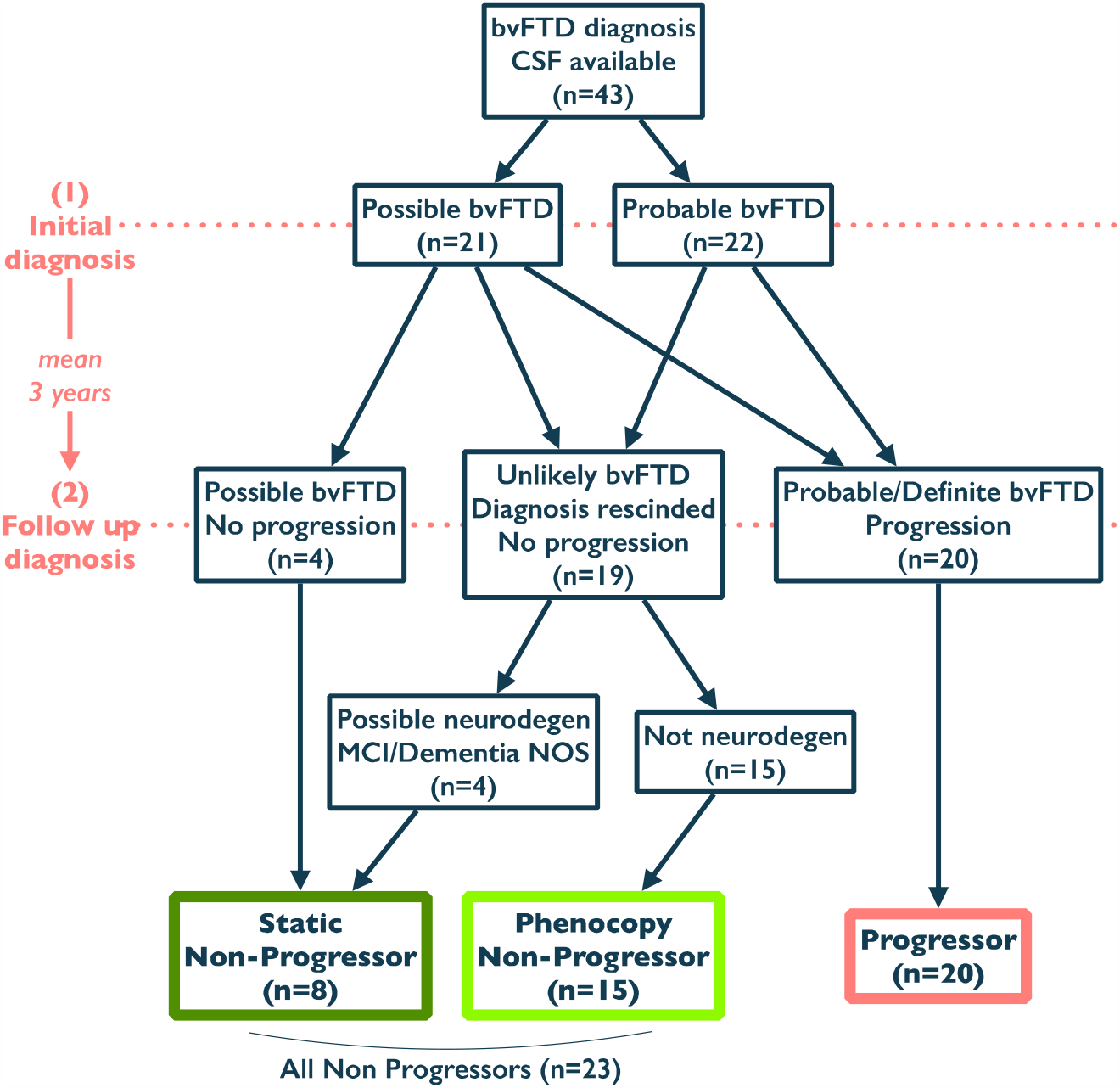
Diagnostic categorisation process. Progressors were patients initially diagnosed with bvFTD, who had progression over time (clinical and/or neuroimaging and/or bedside cognitive/neuropsychological testing, and/or functioning), and retained the bvFTD diagnosis on follow up. Non-Progressors were patients initially diagnosed with bvFTD, but who did not show evidence of progression on follow up (clinical and/or neuroimaging and/or bedside cognitive/neuropsychological testing, and/or functioning). Non-Progressors were further subtyped in to Phenocopy Non-Progressors (the initial bvFTD was eventually completely rescinded in favour of a primary psychiatric/psychological/non-neurodegenerative diagnosis), and Static Non-Progressors (although bvFTD was though less likely, there was still a suspicion of ‘something neurological/neurodegenerative going on’ to explain static cognitive and functional impairments) Final diagnoses for Phenocopy Non-Progressors included: post-traumatic stress disorder, maladaptive personality traits/personality disorders, psychotic disorder, bipolar disorder, subjective cognitive complaints secondary to psychological/psychiatric/substance use issues. Final diagnoses for Static Non-Progressors included: dementia not-otherwise-specified, mild cognitive impairment (vascular, unspecified non-amnestic), mood and personality disorders as potentially contributing factors. bvFTD, behavioural variant frontotemporal dementia; MCI, mild cognitive impairment; NOS, not otherwise specified

Progressors were defined as patients who were initially diagnosed with bvFTD, and retained the bvFTD diagnosis on follow up. In keeping with the retained bvFTD diagnosis on follow up, Progressors had either a) clear progression (clinical and/or neuroimaging and/or bedside cognitive/neuropsychological testing, and/or functioning), and/or b) their diagnostic classification strength progressed (i.e. from possible to probable, possible/probable to definite).

Non-Progressors were patients initially diagnosed with bvFTD, but who did not show evidence of progression on follow up (clinical and/or neuroimaging and/or bedside cognitive/neuropsychological testing, and/or functioning). To explore possible heterogeneity within the Non-Progressor Group, Non-Progressors were further subtyped in to Phenocopy Non-Progressor, and Static Non-Progressor. Phenocopy Non-Progressors were defined as patients where the initial bvFTD diagnosis was eventually completely rescinded, a neurodegenerative disorder was deemed unlikely, and the final diagnosis was of one of a primary psychiatric / psychological / non-neurodegenerative disorder. Static Non-Progressors were patients where although there was no clear evidence of progression, there was a neurological/neurodegenerative cause was still suspected as the cause of static cognitive and functional impairments, and unlike the Phenocopy Non-Progressors, there was no clear primary psychiatric / psychological or non-neurodegenerative disorder diagnosis to fully explain the presentation. Two authors (Eratne, Keem) independently performed file reviews and categorisation. Data extraction and diagnostic categorisation was done blinded to NfL and other biomarker data.

CSF analysis has been previously described [30] [Eratne et al., Alzheimer’s & Dementia, in press, 2021]. Briefly. all participants had CSF collected in a polypropylene tube during their initial assessment, analysed in duplicate for Aβ42, T-tau and P-tau at the National Dementia Diagnostic Laboratory (NDDL), Melbourne, using INNOTEST ELISA (Fujirebio, Ghent, Belgium), remaining samples stored at −80°C. NfL was analysedat NDDL via NF-light ELISA (UmanDiagnostics, Umea, Sweden), according to the manufacturer protocols. Diluted CSF (1+1) and reconstituted standards were added to the plate in duplicate, incubated, washed. Samples displaying concentrations above the highest standard point were further diluted and re-assayed. Two internal controls of pooled CSF were included in each NfL plate. Mean intra-assay coefficient and inter-assay coefficient of variation was 6.2% and 11.3%, respectively.

Healthy control data was available from the ‘Australian Imaging, Biomarker & Lifestyle’ (AIBL) study, previously described [30]. Briefly, criteria for inclusion were: age at time of CSF sampling less than 70; no psychiatric or neurological diagnosis at time of CSF sampling nor in 18 months prior; negative amyloid PET; normal CSF Aβ1-42, T-tau and P-tau levels; classification as healthy control based on formal neuropsychology assessment.

This study was approved by Human Research Ethics Committees at Melbourne Health (2016.038: approval for retrospective biomarker analyses; 2018.371, 2020.142: participants provided consent), and Florey Institute of Neurosciences and Mental Health (1648441.1).

### Statistical analysis

Statistical analysis was undertaken using SPSS 27 and 28. General linear models (GLMs) were performed for comparison of NfL, Aβ1-42, T-tau and P-tau in Progressors versus Non-Progressors, with age as a covariate. ROC curve analyses determined areas under the curve (AUC), Youden’s method to determine optimal cut-off, sensitivity and specificity of biomarkers in distinguishing between groups. GLMs had bias-corrected and accelerated (BCa) confidence intervals (CIs) derived via nonparametric bootstrapping, 1000 replicates. Statistical significance was defined as any CI not capturing the null-hypothesis value (at the 95% level).

## Results

### Study cohort details and clinical variables (Table 1)

Forty-three patients met inclusion criteria: 20 bvFTD Progressors, and 23 Non-Progressors (comprising 15 Phenocopy Non-Progressors, 8 Static Non-Progressors). The mean age at initial assessment, age at CSF collection, age at onset, and proportion of females, were not significantly different between the patient groups. All patient groups had similar, several years of clinical follow up information available after the initial bvFTD diagnosis (mean 3 years). 20 controls were included, mean age 66 years.

Non-Progressors had a higher proportion of possible bvFTD at initial diagnosis, compared to Progressors (78% vs. 15%), and thus a lower proportion of probable bvFTD (22% vs. 85%). Several clinical variables were more frequent in all Non-Progressors, Phenocopy and Static Non-Progressors, compared to Progressors: substance use (approximately double in Non-Progressors), previous psychiatric diagnoses (close to 90% in Non-Progressors, versus 35% in Progressors), and non-psychotic psychiatric symptoms at presentation (over 80% in Non-Progressors, 60% in Progressors). New psychiatric symptoms (defined as within 5 years of presentation) were higher in Non-Progressors, particularly Phenocopy Non-Progressors (80% vs. 20% Progressors). All patients had functional impairment with low Global Assessment of Functioning (GAF) scores, and the majority of patients in all groups had abnormalities on neurological examination. All Progressors had abnormal MRI (and almost all had abnormal SPECT/PET), and the majority of Non-Progressors had abnormalities on neuroimaging (although not always frontotemporal suggestive of bvFTD). All patient groups had similar impairments on the Neuropsychiatry Unit Cognitive Assessment Tool (NUCOG) bedside cognitive screening instrument total score and cognitive subdomains. Although Progressors had slightly poorer performance on the NUCOG total score and executive function domain, this was not statistically different. No patients, regardless of Progressor or Non-Progressor status, had normal performance on formal neuropsychological assessment.

### CSF neurofilament light chain and other biomarkers

NfL concentrations were lower in Non-Progressors (Non-Progressors Mean, M=554pg/mL, 95%CI:[461, 675], Phenocopy Non-Progressors M=459pg/mL, 95%CI:[385, 539], Static Non-Progressors M=730pg/mL, 95%CI:[516, 940]), compared to bvFTD Progressors (M=2397pg/mL, 95%CI:[1607, 3332]) (Table 2 and Figure 2). Although lowest levels and the narrowest range was seen in the Phenocopy Non-Progressor group, levels were not statistically different to Static Non-Progressors (GLM mean difference, Mdiff=192, 95%CI:[124, 518]). The differences in NfL levels between Progressors and other groups were large: Progressors versus all Non-Progressors (GLM mean difference, Mdiff=1817, 95%CI:[988, 2745]), versus Phenocopy Non-Progressors (MDiff=1885, 95%CI:[1093, 2721]), versus Static Non-Progressors (Mdiff=1694, 95%CI:[900, 2545]), and versus controls (Mdiff=2097, 95%CI:[982, 3290]).

**Table 2.**
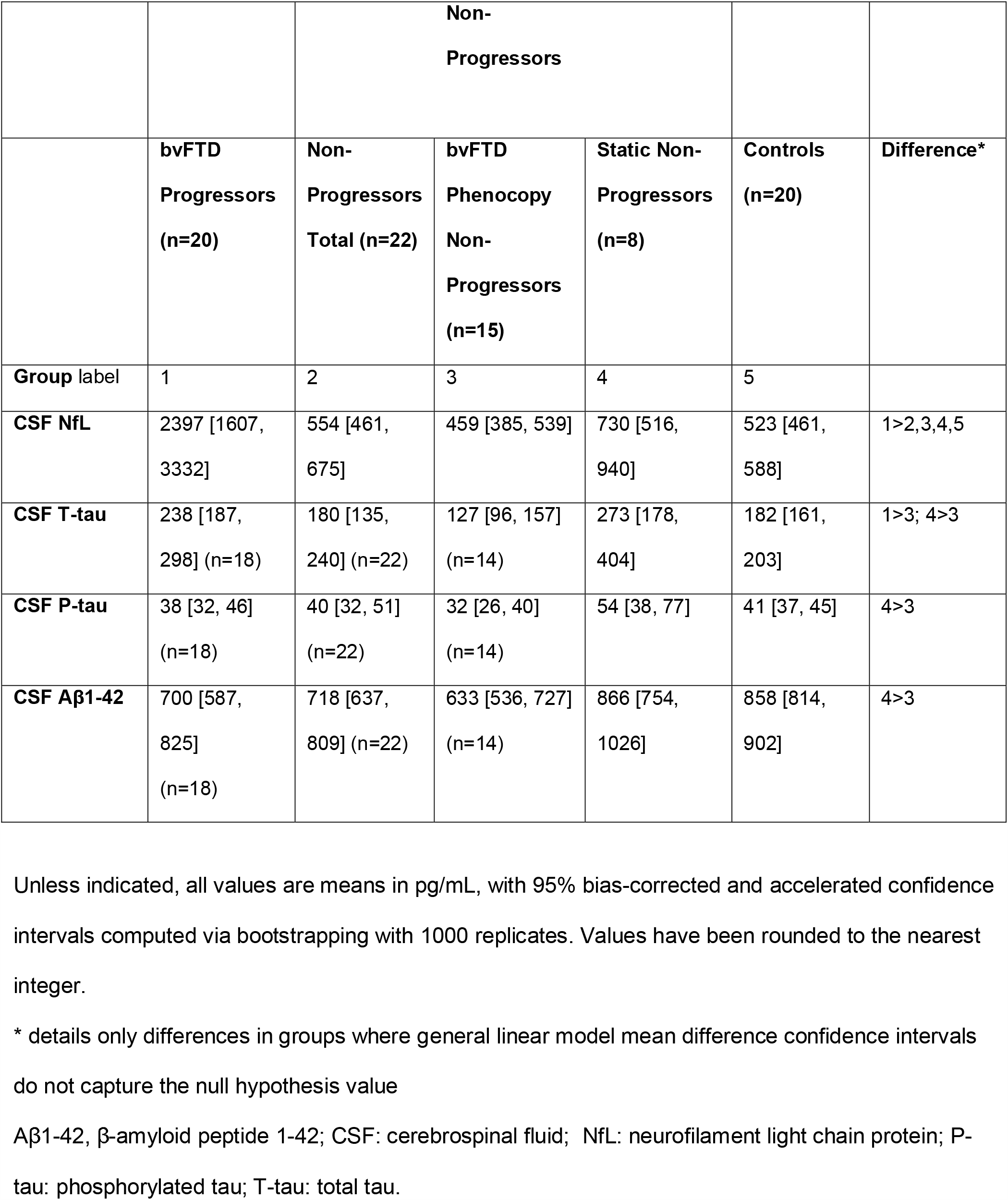
Cerebrospinal fluid biomarker concentrations in bvFTD Progressors, Non-Progressors, and controls.

|  |  | **Non-Progressors** |  |  |  |  |
| --- | --- | --- | --- | --- | --- | --- |
|  | **bvFTD Progressors (n=20)** | **Non-Progressors Total (n=22)** | **bvFTD Phenocopy Non-Progressors (n=15)** | **Static Non-Progressors (n=8)** | **Controls (n=20)** | **Difference\*** |
| **Group label** | 1 | 2 | 3 | 4 | 5 |  |
| **CSF NfL** | 2397 [1607, 3332] | 554 [461, 675] | 459 [385, 539] | 730 [516, 940] | 523 [461, 588] | 1>2,3,4,5 |
| **CSF T-tau** | 238 [187, 298] (n=18) | 180 [135, 240] (n=22) | 127 [96, 157] (n=14) | 273 [178, 404] | 182 [161, 203] | 1>3; 4>3 |
| **CSF P-tau** | 38 [32, 46] (n=18) | 40 [32, 51] (n=22) | 32 [26, 40] (n=14) | 54 [38, 77] | 41 [37, 45] | 4>3 |
| **CSF Aβ1-42** | 700 [587, 825] (n=18) | 718 [637, 809] (n=22) | 633 [536, 727] (n=14) | 866 [754, 1026] | 858 [814, 902] | 4>3 |

Unless indicated, all values are means in pg/mL, with 95% bias-corrected and accelerated confidence intervals computed via bootstrapping with 1000 replicates. Values have been rounded to the nearest integer.
\* details only differences in groups where general linear model mean difference confidence intervals do not capture the null hypothesis value
Aβ1-42, β-amyloid peptide 1-42; CSF: cerebrospinal fluid; NfL: neurofilament light chain protein; P-tau: phosphorylated tau; T-tau: total tau.

**Figure 2.**
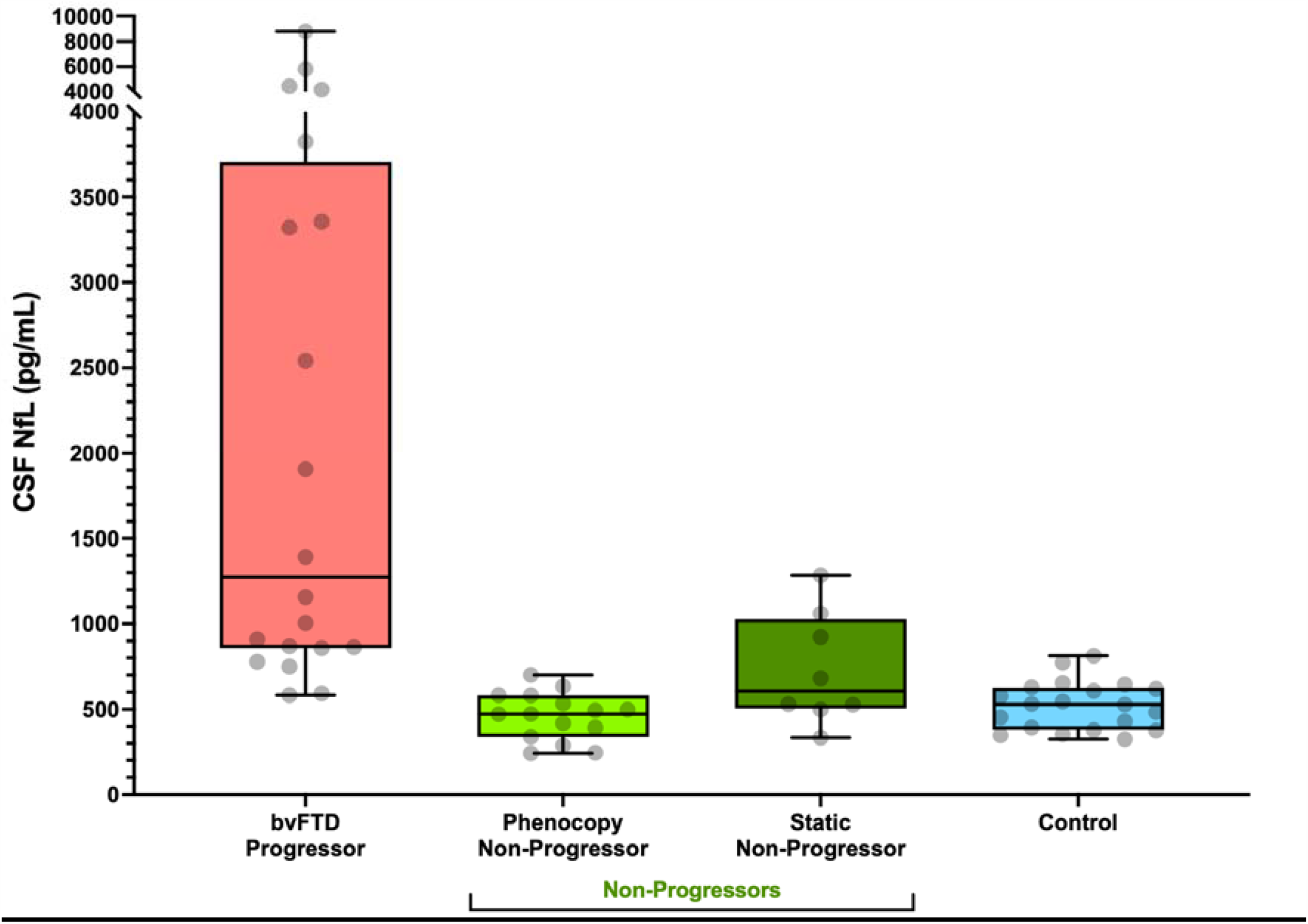
Box plot of cerebrospinal fluid (CSF) neurofilament light (NfL) between different groups. bvFTD, behavioural variant frontotemporal dementia; CSF, cerebrospinal fluid; NfL, neurofilament light.

T-tau levels were higher in Progressors (M=238pg/mL, 95%CI:[187, 298]) compared to Phenocopy Non-Progressors (M=127pg/mL, 95%CI:[96, 157], Mdiff=108, 95%CI:[44, 176]), but not Static Non-Progressors (M=273pg/mL, 95%CI:[178, 404]). Static Non-Progressors had higher T-tau levels than Phenocopy Non-Progressors (Mdiff=139, 95%CI:[26, 298]), Table 2, Supplementary Figure 1.

Static Non-Progressors also had higher P-tau levels than Phenocopy Non-Progressors (Mdiff=21, 95%CI:[3, 42]), Table 2, Supplementary Figure 2. Aβ42 levels were lower in Phenocopy Non-Progressors compared to Static Non-Progressors (Mdiff=218, 95%CI:[77, 363]) and controls (Mdiff=170, 95%CI:[18, 300]), Table 2, Supplementary Figure 3. There were no sex differences in any biomarker levels.

### NfL, T-tau, P-tau and Aβ1-42 as diagnostic tests

NfL distinguished bvFTD Progressors from all Non-Progressors with the highest accuracy (area under the curve, AUC: 0.92, 90% sensitivity, 87% specificity at optimal cut-off of 726pg/mL), superior to T-tau (AUC 0.69, AUC difference 0.23, 95%CI:[0.09, 0.38], 72% sensitivity, 73% specificity, cut-off 192pg/mL), Figure 3 and Supplementary Material. NfL at the 726pg/mL cut-off distinguished ND from PSY with 86% positive predictive value (PPV), 91% negative predictive value (NPV), 6.8 positive likelihood ratio (LR+ve), 0.11 negative likely ratio (LR-ve), accurately classifying 88% (38/43) of patients.

**Figure 3.**
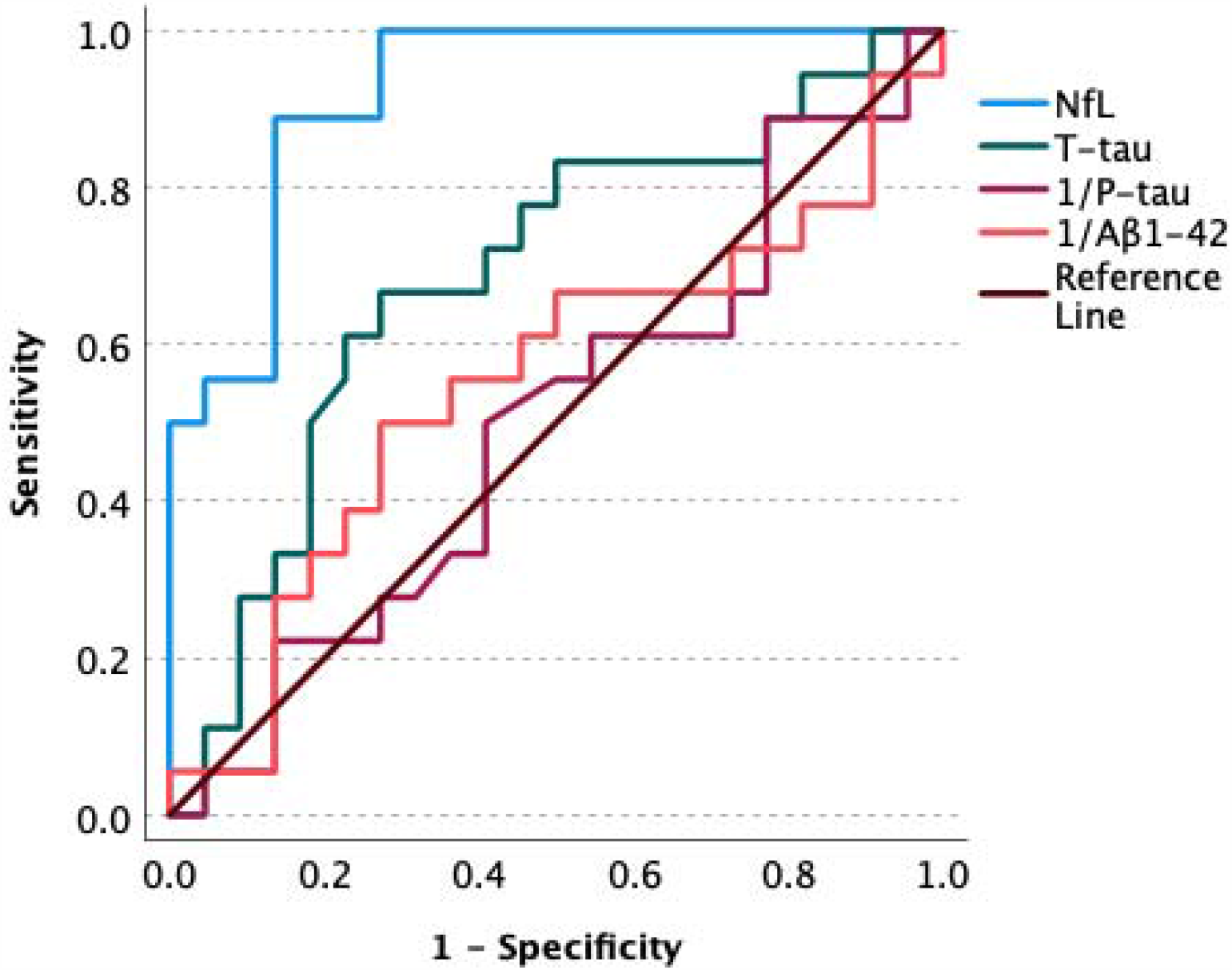
ROC Curve for behavioural variant frontotemporal dementia Progressors versus all Non-Progressors (comprising Phenocopy Non-Progressors and Static Non-Progressors) ROC: receiver operating characteristic; NfL, neurofilament light; Aβ1-42, β-amyloid peptide 1-42; P-tau, phosphorylated tau; T-tau, total tau.

NfL performed particularly well in distinguishing Progressors from Phenocopy Non-Progressors (AUC 0.99, 90% sensitivity, 100% specificity at cut-off of 726pg/mL, and at a cut-off of 583pg/mL, 100% sensitivity, 87% specificity, 100% PPV, 88% NPV, 0.10 LR-ve, 94% accuracy). NfL was once again superior to T-tau for this distinction (AUC difference 0.18, 95%CI:[0.02, 0.33]).

NfL had lower specificity in distinguishing Progressors from Static Non-Progressors (AUC 0.82, 90% sensitivity, 63% specificity, cut-off 716pg/mL). Combining NfL and Aβ42 in to an NfL:Aβ42 ratio resulted in improved AUC and specificity (AUC 0.88, 73% sensitivity, 100% specificity, cut-off 1.40), although this was not statistically significant (AUC difference 0.06, 95%CI:[-0.24, 0.11]). More information on these other comparisons are available in Supplementary Material.

## Discussion

Our study found that baseline CSF NfL levels differentiated bvFTD from non-progressive ‘phenocopies’, with high accuracy, in a cohort of patients from a real-world clinical setting. Our findings have important clinical translation implications, supporting the use of CSF NfL, at initial assessment, as a sensitive and specific biomarker in predicting and distinguishing, progressive bvFTD from non-progressive and mimicking syndromes that have historically been subsumed under the ‘phenocopy bvFTD’ label. This is a common, challenging clinical dilemma, with patients often needing several years and multiple assessments and investigations before a phenocopy ‘non-progressor’ diagnosis can more confidently be made.

A novel aspect of our study was exploring Non-Progressors in more detail, given the likely heterogeneity within this group. We sub-categorised non-progressors in to ‘Phenocopy Non-Progressors’, where a primary psychiatric/psychological non-neurodegenerative formulation was ultimately made, compared to ‘Static Non-Progressors’, where there was still clinical suspicion of a neurological/neurodegenerative cause for static cognitive and functional impairments. There has been speculation that this group may still have neurodegenerative aetiology, subclinical, or at an extreme slowly progressive end of a bvFTD spectrum. Our findings in the Static Non-Progressor group, of slightly higher NfL, and in particular higher total tau levels, compared to the Phenocopy Non-Progressors, may be consistent with a subclinical bvFTD or another slowly progressive neurodegenerative or tau-related syndrome, albeit at a very low rate that is not demonstrated clearly on serial clinical and neuroimaging assessments. Interestingly, it was the Phenocopy Non-Progressor group that had lower AB42 levels, similar to levels seen in the bvFTD progressors. However, given the small number of Static Non-Progressors, caution is required in interpreting these exploratory findings. To help understand the possible heterogeneity in apparent non-progressors, further research in larger cohorts, with serial NfL levels, detailed clinical, biomarker, and genetic phenotyping, is warranted.

Our findings of overlap on a range of clinical variables at baseline, adds further weight to how challenging and complex the distinction between Progressors and Non-Progressors is, in real-world clinical practice. Bedside cognitive testing did not discriminate Progressors from Non-Progressors, and bvFTD Progressors had memory as well as executive function impairments. All Non-Progressors had impairments on neuropsychological assessment, and most Non-Progressors had abnormalities on structural and functional neuroimaging. These results, and our finding that Non-Progressors were more likely to only initially meet criteria for possible bvFTD, support other evidence on the greater uncertainty and diagnostic instability of possible bvFTD compared to probable/definite, and recommendations to be cautious about a diagnosis of bvFTD in the absence of neuroimaging changes consistent with bvFTD [11,14,18]. However, a significant proportion of Phenocopy Non-Progressors (33%, 5/15) had frontotemporal imaging changes that led to initially meeting criteria for probable bvFTD. Our findings would support increased suspicion of Non-Progressor in patients with a previous psychiatric history, and where psychiatric symptoms (separate to behavioural change) are more prominent features at presentation. However, while there some differences at group levels, similar to what has been described in the literature [11,13,15,39], given the complexity and overlap between patient groups, no single clinical variable nor combination variables separated Progressors from Non-Progressors, as well as a single biomarker, NfL.

Our data supports an emerging consensus in the literature [10] that bvFTD phenocopies – represented predominately by our Phenocopy Non-Progressors – may be considered a ‘pseudodementia’ syndrome with several common factors: male sex, personality vulnerabilities, significant social stressors that may injure self-identity, maladaptive coping strategies including significant substance misuse, chronic dysthymia or ‘reactive’ depression, and for some, the additional development of melancholic depressive features to constitute a ‘double depression’. The majority of Non-Progressors in our cohort had a longstanding history of psychiatric symptoms (most commonly both non-melancholic/’reactive’ and melancholic depressive symptoms), psychiatric history (typically major depressive disorder, PTSD), and Cluster B (especially borderline, histrionic and narcissistic) and Cluster C (primarily dependent and avoidant) personality constructs. Non-Progressors were more likely to have additional, new/superimposed symptoms, on this background, manifesting in context of interpersonal and psychosocial difficulties. In contrast, Static Non-Progressors did not have elevated rates of new psychiatric symptoms, but did have high rates of previous psychiatric diagnoses, and psychiatric symptoms at presentation, similar to Phenocopy Non-Progressors.

While this study is limited by its relatively small cohort size, our total case numbers were superior or comparable to several other retrospective cohort studies [13,14,16,17], and contribute to the limited literature in this area, in particular regarding biomarkers. The retrospective design is an inherent limitation, mitigated by the consistency of multidisciplinary and multimodal assessments within a single tertiary service, and rigorous file review blinded to biomarkers and conducted by two investigators separately. Although our cohort had several years of follow up and serial, gold-standard multidisciplinary and multimodal assessments and diagnosis based on established diagnostic criteria, it is possible that longer follow up could have revealed more insights, and most patients lacked definitive genetic or pathological confirmation. We did not have phonemic fluency data on participants, which is a limitation of the NUCOG and our dataset, given other studies have found discriminatory potential [39]. The complexity of those patients with multifaceted symptomatology, comorbidities and psychiatric histories included in this study that warranted referral to a tertiary neuropsychiatry service for assessment and diagnosis, may not necessarily be generalisable to other services. However, most such patients are seen within less sub-specialised services nor do most patients have access to such tertiary services, and in addition the broad inclusion criteria of this study helped mitigate this possibility, and is a key strength relative to much of the literature.

This study demonstrated the diagnostic utility of CSF NfL in distinguishing progressive bvFTD from non-progressive variants and mimics, at baseline, with high accuracy. As blood NfL correlates strongly with CSF levels [25,40], a simple blood test as a first-tier test at initial assessment could strongly support the diagnosis of progressive bvFTD, versus phenocopy or mimicking syndromes. This would be of immense benefit in primary and secondary, and rural healthcare settings where the resources and expertise required to reach a gold-standard consensus diagnosis of bvFTD are often unavailable. With further study, a speculative diagnostic algorithm incorporating CSF NfL, other biomarkers, and clinical variables, could be developed to not only accurately differentiate bvFTD from non-progressive variants, but further delineate truly non-neurodegenerative from other aetiologies (Supplementary Material). The capacity to accurately diagnose a progressive neurodegenerative illness and identify a potentially reversible condition mimicking bvFTD at time of initial assessment, could dramatically reduce delay and misdiagnosis, and improve outcomes for patients, their families, and healthcare systems. NfL, supplemented by T-tau and Aβ1-42, promises to transform complex diagnosis and prognostication in the highly vulnerable cohort of patients presenting with frontotemporal neuropsychiatric impairments, and building on these findings, larger prospective studies assessing CSF and plasma biomarkers, are underway.

## Data Availability

All data produced in the present study are available upon reasonable request to the authors

## ACKNOWLEDGEMENTS AND FUNDING SOURCES

We are grateful for funding that supported this work: the Trisno Family Research Grant in Old Age Psychiatry, three NorthWestern Mental Health Research Seed Grants, MACH MRFF RART 2.1, and NHMRC (1185180). Finally, the authors would like to thank all the patients and their families for their participation.

The corresponding author had full access to all the data in the study and had final responsibility for the decision to submit for publication.

## APPENDIX 1 Collaborators

### Collaborators associated with The MiND Study Group

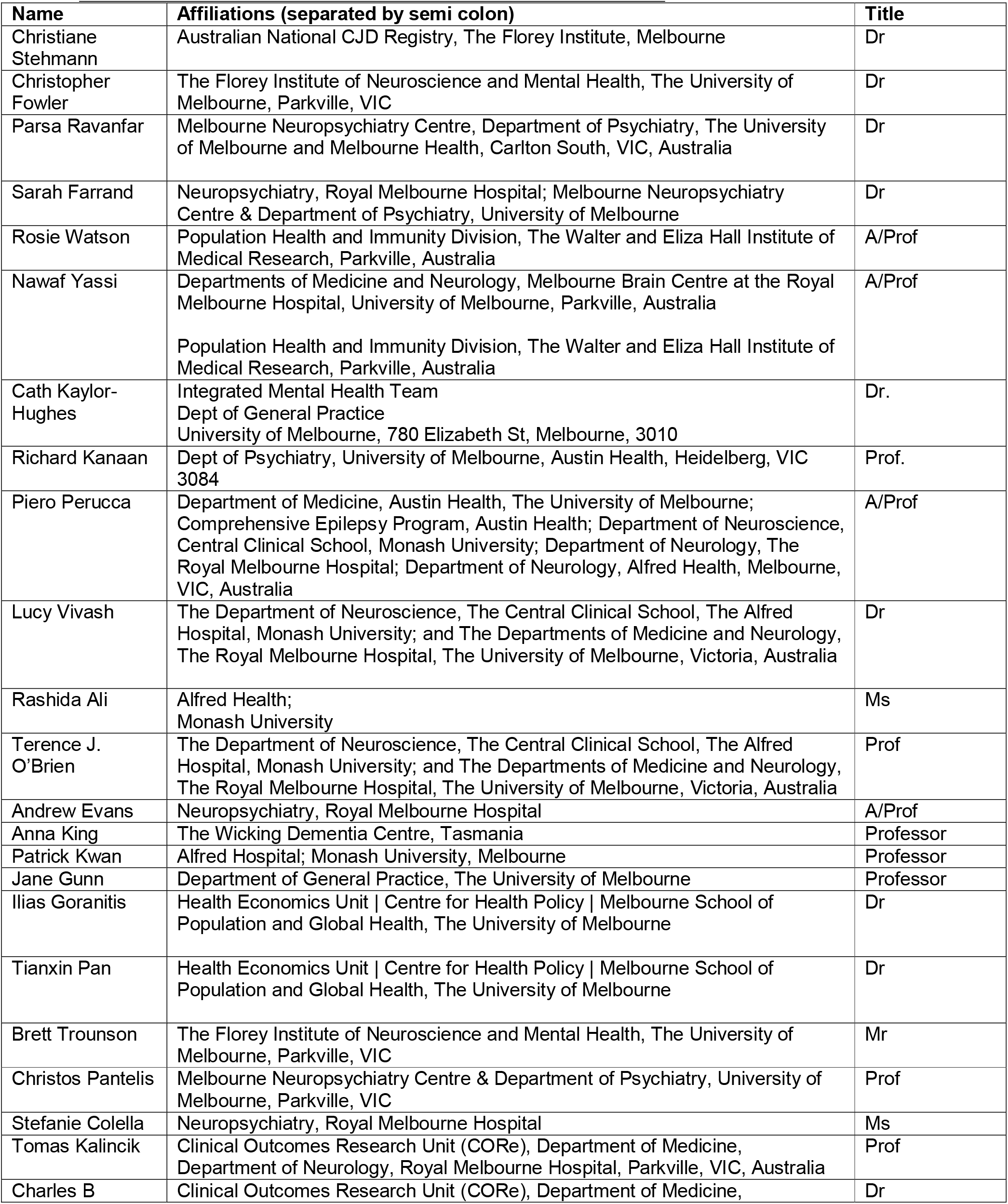
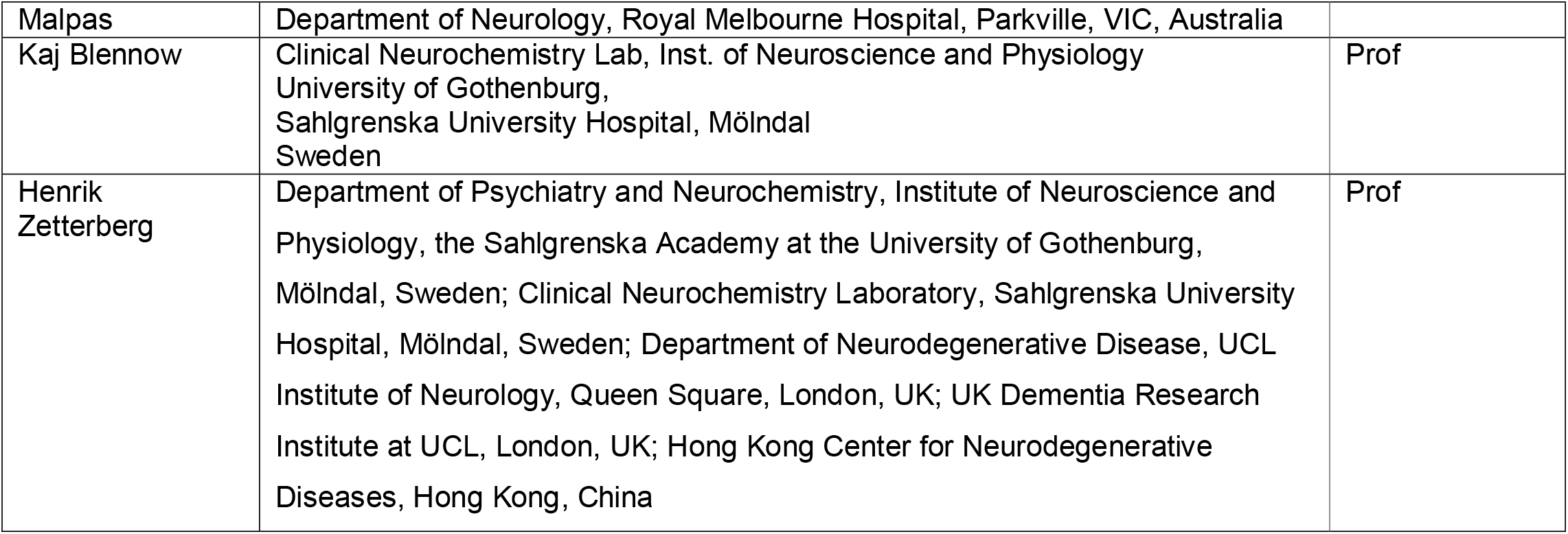

**Supplementary Material (see separate document)**

## Notes

### Competing Interest Statement

The authors have declared no competing interest.

### Author Declarations

Ethics committees of The Royal Melbourne Hospital and The Florey Institute of Neuroscience and Mental Health gave ethical approval for this work. This study was approved by Human Research Ethics Committees at The Royal Melbourne Hospital/Melbourne Health (2016.038: approval for retrospective biomarker analyses; 2018.371, 2020.142: participants provided consent), and Florey Institute of Neurosciences and Mental Health (1648441.1).

