## Supplementary Material for "Cerebrospinal fluid neurofilament light chain differentiates behavioural variant frontotemporal dementia progressors from ‘phenocopy’ non-progressors"

**Supplementary Material (see separate document)**

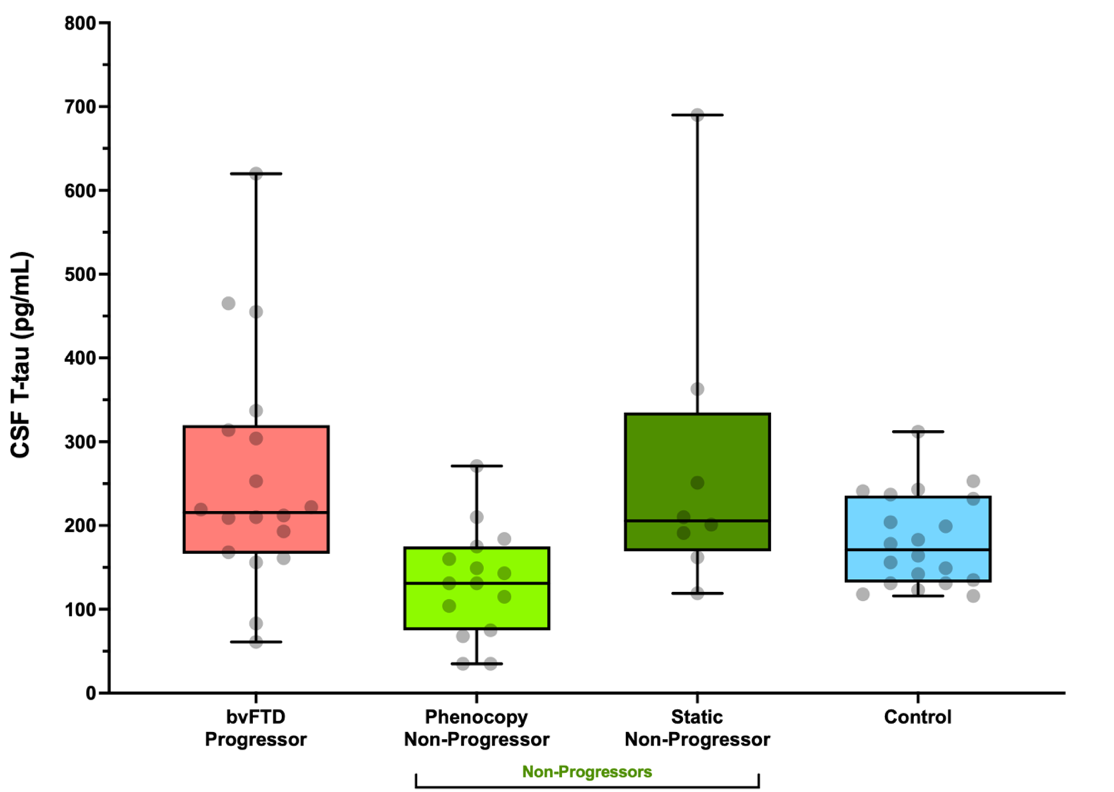

Supplementary Figure 1. Cerebrospinal fluid total tau in the different groups

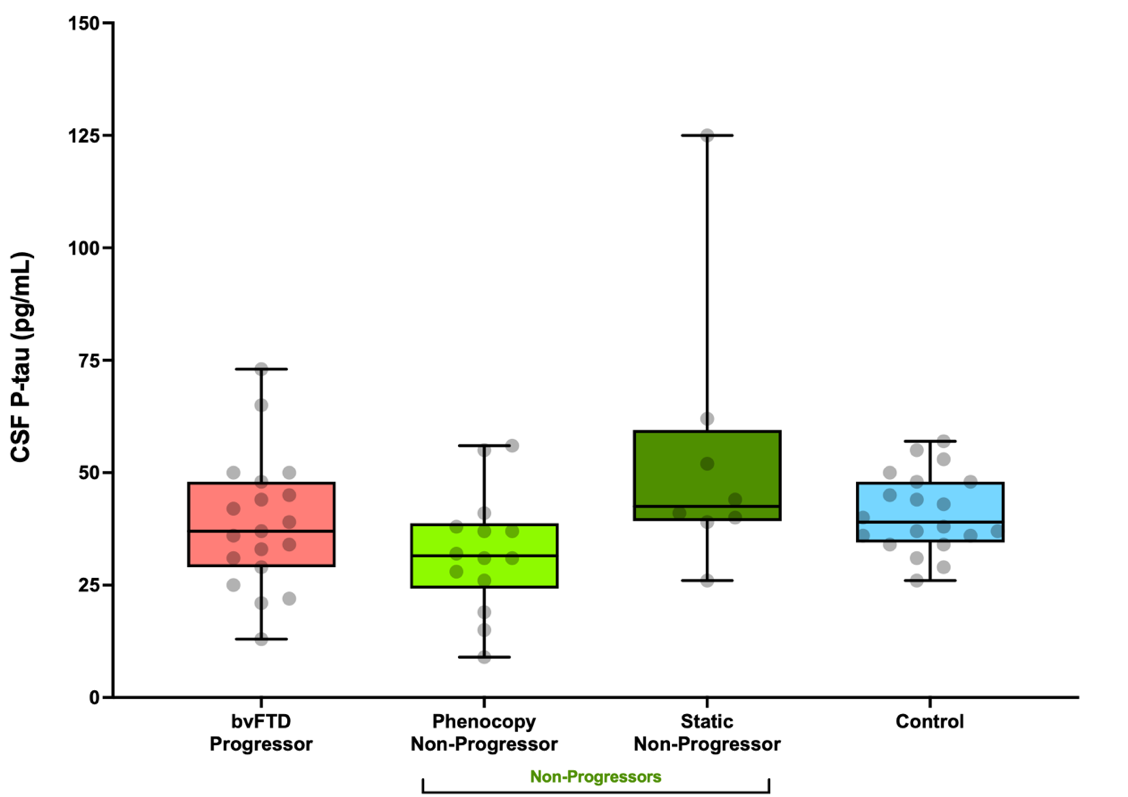

Supplementary Figure 2. Cerebrospinal fluid phosphorylated tau in the different groups

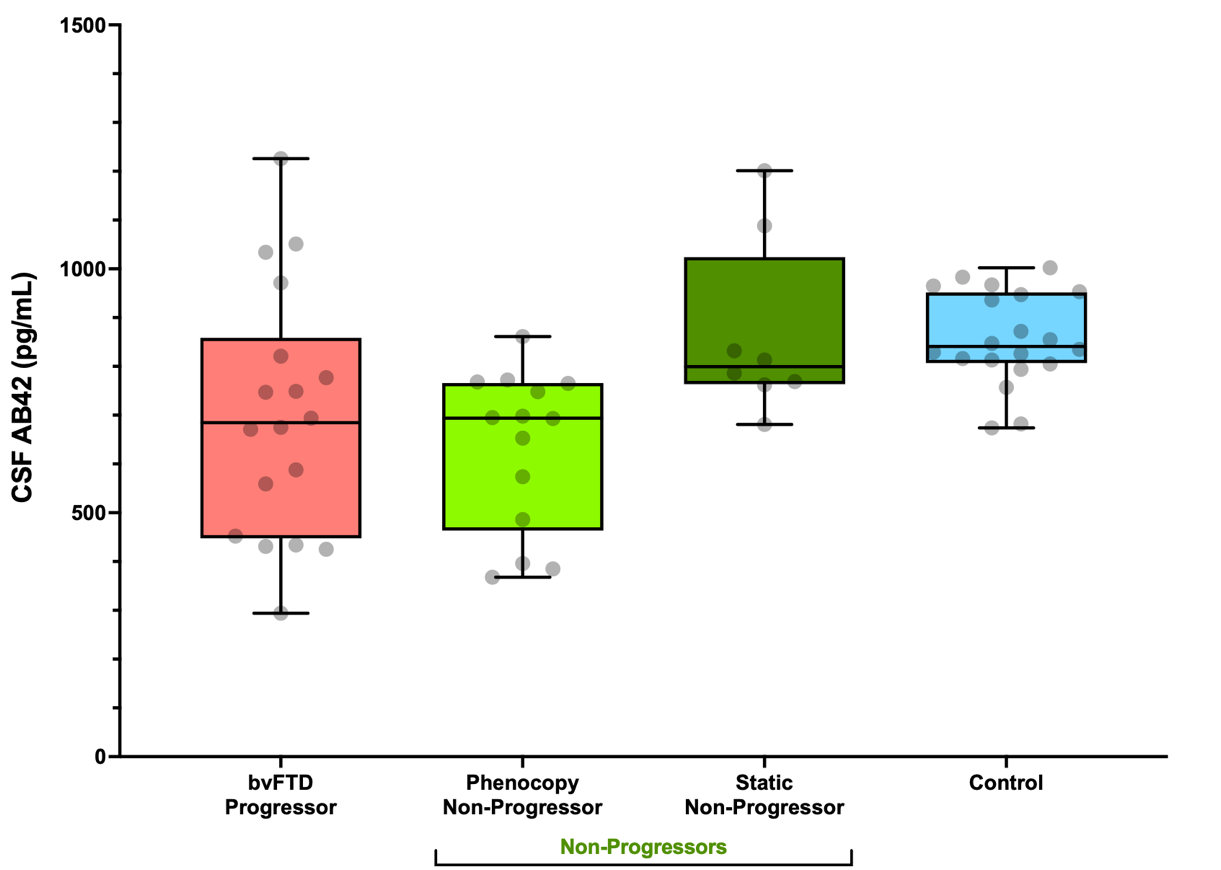

Supplementary Figure 3. Cerebrospinal fluid amyloid beta 1-42 in the different groups

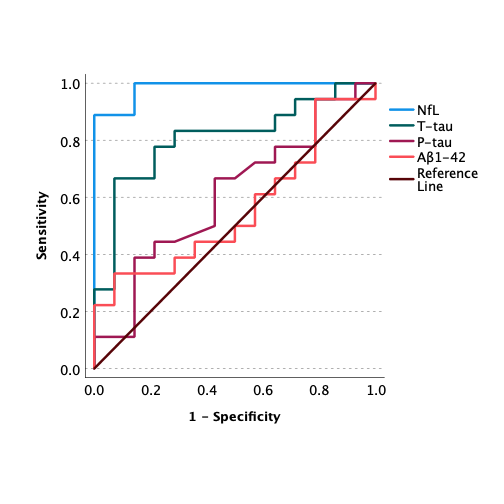

Supplementary Figure 4. ROC curve for behavioural variant frontotemporal dementia Progressors versus Phenocopy Non-Progressors

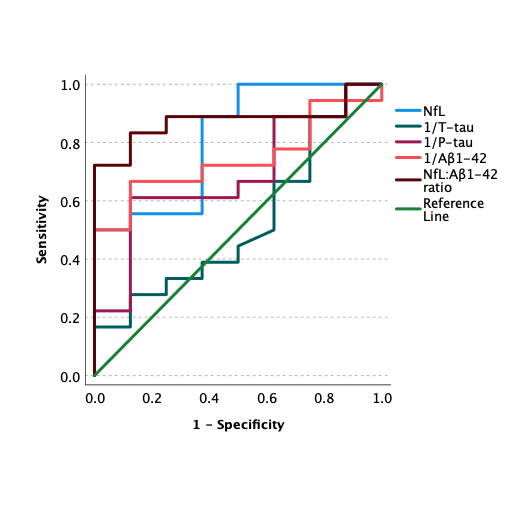

Supplementary Figure 5. ROC curve for behavioural variant frontotemporal dementia Progressors versus Static Non-Progressors

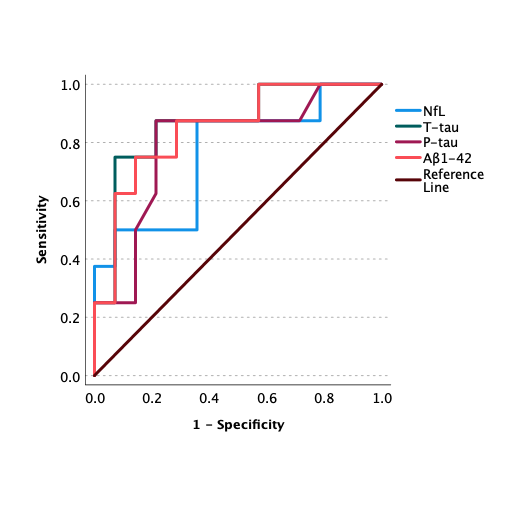

Supplementary Figure 5. ROC curve for Static Non-Progressors vs Phenocopy Non-Progressors

|  | CSF biomarker | AUC [95% CIs] | Cut-off (pg/mL) | Sensitivity | Specificity |
| --- | --- | --- | --- | --- | --- |
| Progressors vs. All Non-Progressors | NfL | 0.92 [0.84, 1.0] | 726 | 90 | 87 |
|  | T-tau | 0.69 [0.52, 0.86] | 192 | 72 | 73 |
|  | P-tau | 0.50 [0.32, 0.68] | - | - | - |
|  | Aβ1-42 | 0.55 [0.36, 0.74] | - | - | - |
| Progressors vs Phenocopy Non-Progressors | NfL | 0.99 [0.96, 1.00] | 726  (583) | 90  (100) | 100  (87) |
|  | T-tau | 0.81 [0.66, 0.96] | 189 | 72 | 87 |
|  | P-tau | 0.61 [0.41, 0.81] ^NS^ | - | - | - |
|  | Aβ1-42 | 0.56 [0.36, 0.76] ^NS^ | - | - | - |
| Progressors vs Static Non-Progressors | NfL | 0.82 [0.64, 0.99] | 716 | 90 | 63 |
|  | T-tau | 0.52 [0.25, 0.78] ^NS^ | - | - | - |
|  | P-tau | 0.65 [0.41, 0.89]^NS^ | - | - | - |
|  | Aβ1-42 | 0.74 [0.55, 0.93] | 769 (less than) | 67 | 88 |
|  | NfL:Aβ1-42 ratio | 0.88 [0.74, 1.00] | 1.40 | 73 | 100 |
| Static Non-Progressors vs Phenocopy Non-Progressors | NfL | 0.77 [0.55, 0.98] | 498 | 88 | 67 |
|  | T-tau | 0.87 [0.70, 1.00] | 187 | 75 | 93 |
|  | P-tau | 0.80 [0.58, 1.00] | 39 | 88 | 79 |
|  | Aβ1-42 | 0.85 [0.68, 1.00] | 768 | 75 | 86 |

Supplementary Table 1. Receiver operating characteristic (ROC) curve analyses for bvFTD Progressors vs. Non-Progressors

AUC: area under the curve; CIs: confidence intervals; NfL: neurofilament light; Aβ1-42, β-amyloid peptide 1-42; P-tau: phosphorylated tau; T-tau: total tau.

NS: Values that captured the null hypothesis value, thus not reaching statistical significance.

| NfL cut-off 726pg/mL | | | NfL cut-off 726pg/mL | | |
| --- | --- | --- | --- | --- | --- |
| A | Progressor | Non-Progressor | B | Progressor | Phenocopy Non-Progressor |
| Above cut-off ‘test positive’ | 18 | 3 | Above cut-off ‘test positive’ | 18 | 0 |
| Below cut-off ‘test negative | 2 | 20 | Below cut-off ‘test negative | 2 | 15 |

**Supplementary Table 2. Crosstabs for neurofilament light chain distinguishing Progressors from All Non-Progressors (A), and Progressors from Phenocopy Non-Progressors (B)**

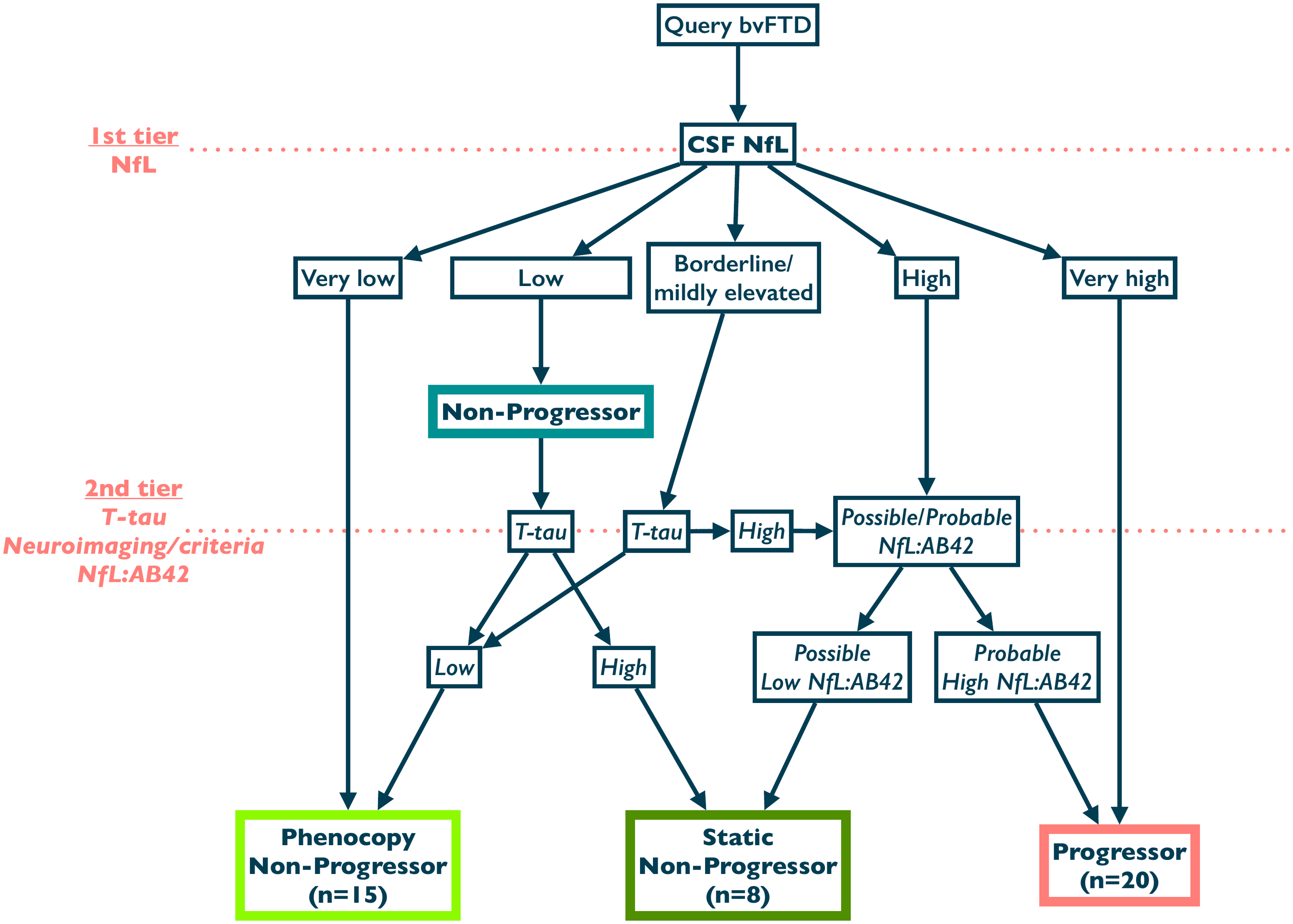

**Supplementary Figure 6. A speculative algorithmic process incorporating cerebrospinal fluid biomarkers and neuroimaging features**

As a first-tier test, a very low CSF NfL (<334pg/mL in our study) would suggest a bvFTD Phenocopy Non-Progressor, while a very high level (>1285pg/mL in our study) would suggest a Progressor. A low level (334-582pg/mL) would be consistent with a Non-Progressor, with T-tau levels helping to differentiate Phenocopy Non-Progressors from Static Non-Progressors. Patients with borderline (583-700pg/mL) levels could benefit from T-tau and NfL:AB42, as well as the presence or absence of frontotemporal neuroimaging abnormalities required to meet diagnostic criteria for probable bvFTD, to differentiate between Phenocopy Non-Progressors, Static Non-Progressors, and Progressors. Patients with high NfL levels (701-1285pg/mL in our study) but only meeting criteria for possible bvFTD, could support a Static Non-Progressor diagnosis over a Progressor.
